# Adaptive deep brain stimulation timed to gait phase improves walking in Parkinson’s disease

**DOI:** 10.1101/2025.08.19.25333759

**Authors:** Kenneth H. Louie, Jannine P. Balakid, Jessica E. Bath, Seongmi Song, Hamid Fekri Azgomi, Jacob H. Marks, Julia T. Choi, Philip A. Starr, Doris D. Wang

## Abstract

Gait dysfunction in Parkinson’s disease (PD) is a major source of disability and is often resistant to traditional deep brain stimulation (DBS). Here, we report a novel neuromodulation paradigm, gait-phase-synchronized adaptive DBS (aDBS), that dynamically modulates stimulation amplitude during contralateral leg swing. In five individuals with PD, we identified personalized neural biomarkers of gait phase from cortical and pallidal field potentials and embedded them into a chronically implanted bidirectional neurostimulator. These biomarkers, derived via a data-driven search, enabled real-time detection of swing phase and sub-second modulation of stimulation amplitude. Acute in-clinic testing showed that aDBS significantly reduced gait variability and improved bilateral symmetry compared to clinically optimized continuous DBS. In a double-blinded, multi-day crossover study, gait-phase-synchronized aDBS was well-tolerated, maintained general motor symptom control, and reduced falls and improved other gait metrics. These findings establish the feasibility of biomarker-driven, movement-synchronized neuromodulation and offer a promising strategy to restore dynamic motor control in PD.

## Main

Gait disturbances in people with Parkinson’s disease (PD) are among the most debilitating symptoms, reducing quality of life, increasing the risk of falls, and diminishing independence^1–3^. Although deep brain stimulation (DBS) therapy may initially improve gait disturbances, its efficacy often diminishes over time^4–10^. Previous attempts to enhance gait outcomes with DBS, such as lowering stimulation frequency^11–18^ or changing stimulation contact location^19–21^ have produced mixed results. Improvements in gait often come at the cost of worsened tremor control^18,22^ or a reduction in falls accompanied by increased gait variability^23^. Thus, a novel stimulation approach is needed to improve gait without compromising the treatment of other PD symptoms.

Current DBS approaches may fail to address gait disturbances because gait is a complex and dynamic motor behavior requiring the precise coordination of muscle contractions and relaxations across all limbs. During locomotion, the human body must maintain balance while each leg alternates between the stance phase, when the foot contacts the ground, and the swing phase, when the foot is in the air. However, current DBS therapy typically delivers continuous stimulation without adjusting to patients’ movement states, which can exacerbate gait impairments^24,25^.

Recent studies have demonstrated that local field potential (LFP) spectral power within deep brain nuclei fluctuates naturally throughout the day, influenced by sleep^26^, mood^27^, and medication states^28–30^. During “on” medication states, when dopaminergic therapy effectively controls motor symptoms, continuous DBS may cause overstimulation, leading to hyperkinetic symptoms such as dyskinesia. Similar state-dependent LFP fluctuations have been observed during gait^31–37^, with distinct increases and decreases in power corresponding to different phases of the gait cycle. Delivering continuous stimulation without accounting for these cyclical neural dynamics may disrupt the natural synchronization required for efficient locomotion and worsen gait performance.

To address these fluctuations, adaptive DBS (aDBS)—an approach that adjusts stimulation parameters based on a real-time control signal related to symptom severity—has gained increasing interest^38,39^. aDBS has been shown to effectively treat dyskinesia^40^ and bradykinesia/rigidity^41–44^, performing as well as or better than continuous DBS, while remaining safe and tolerable. The success of aDBS in managing other PD symptoms suggests that it may also be effective for improving gait disturbances. We hypothesized that synchronizing basal ganglia stimulation to the natural gait cycle could enhance gait patterns in PD by allowing physiological synchronization to occur while selectively suppressing pathological signals.

To test this hypothesis, we developed an aDBS therapy that rapidly modulates stimulation amplitude during contralateral leg swing in five individuals with PD and gait dysfunctions (Fig. 1a). We investigated the pallidum and motor cortical areas for evidence of gait-phase modulation and developed a data-driven approach to identify patient-specific biomarkers of contralateral leg swing in each hemisphere. During free overground walking, patients exhibited unique frequency bands and recording sites that captured spectral power changes during contralateral swing. Acute in-clinic application of aDBS reduced step length and step time variability and improved gait symmetry compared to clinically optimized continuous DBS (cDBS). Importantly, in double-blinded trial testing continuous and two different gait-phase synchronized aDBS settings over longer term at home, patients not only tolerated rapid stimulation adjustments, but also maintained general PD symptom control, and exhibited multiple improved gait metrics. Together, these findings demonstrate the feasibility of rapid, gait-phase-synchronized aDBS for improving gait in PD, a promising novel therapy for treating advanced gait disorders in PD.

**Figure 1.**
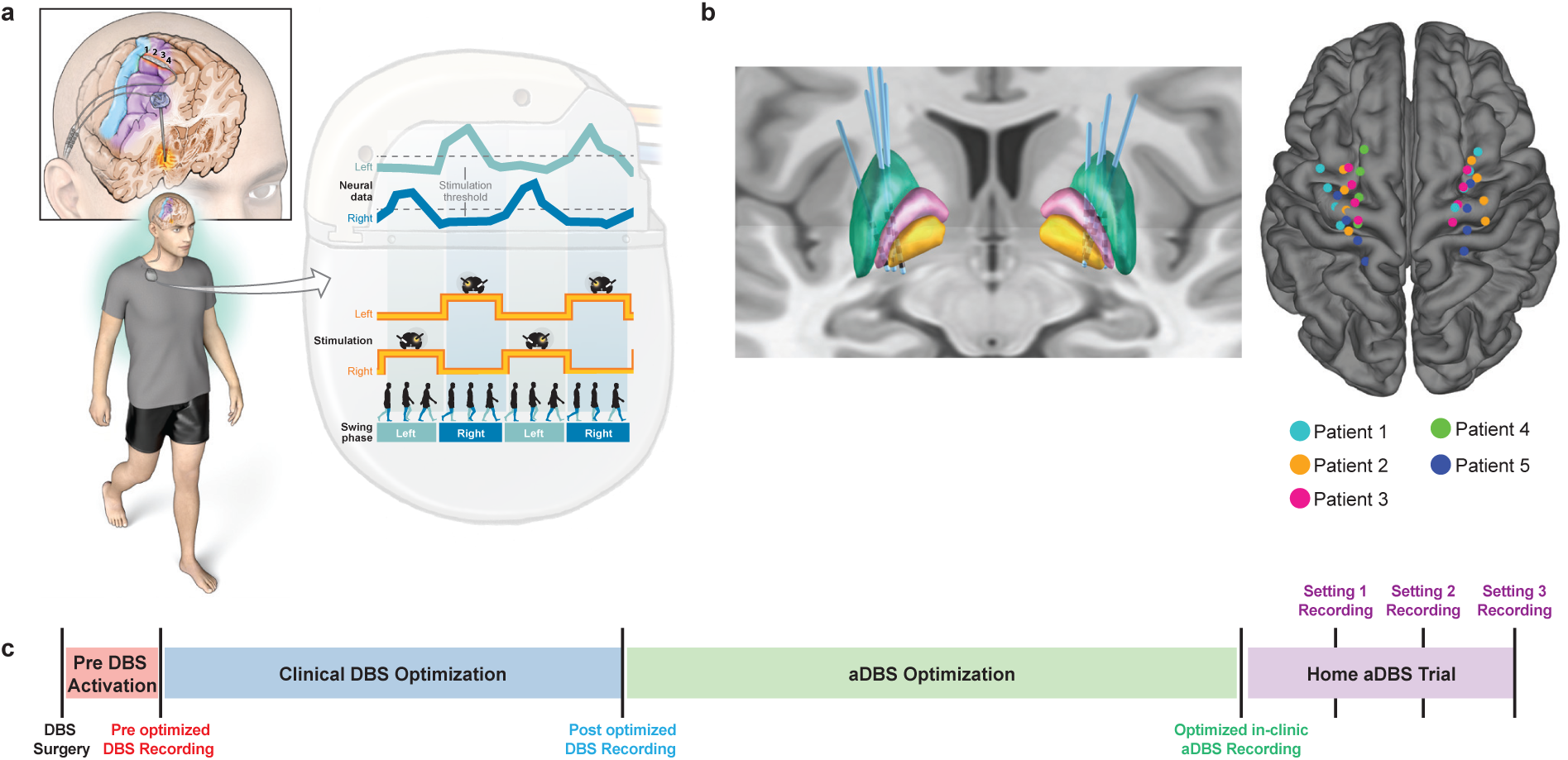
Overview of adaptive DBS system and study design. **a,** Adaptive DBS (aDBS) control strategy for independent, phase-specific modulation of stimulation amplitude in each hemisphere. Neural signals were continuously recorded from the internal segment of the globus pallidus (GPi) or cortex (teal and blue traces), and spectral power within a specified frequency band was computed. When spectral power crossed a defined threshold (dashed gray line), stimulation amplitude (yellow trace) increased to the clinically optimized value during contralateral leg swing and decreased to 0.5x the optimized amplitude during other gait phases. **b,** Reconstruction of DBS leads and cortical electrode strips, normalized to Montreal Neurological Institute (MNI) space. Cortical electrodes were overlaid on a standard brain atlas. The internal pallidum is shown in orange, external pallidum in pink, and striatum in green. **c,** Study timeline divided into four phases. Phase 1: postoperative period before DBS activation (red). Phase 2: standard clinical optimization of DBS parameters (blue). Phase 3: testing and optimization of adaptive settings (green). Phase 4: home testing of optimized aDBS settings in three patients (Patient 2–4) under double-blind conditions. Three stimulation settings were tested: aDBS ramp-up (0.5x to 1.0x amplitude) during contralateral leg swing, aDBS ramp-down (1.0x to 0.5x amplitude) during contralateral leg swing, and clinically optimized continuous DBS.

## Results

### Patients overview and study design

Patients were recruited from a cohort of individuals with PD with gait problems who were eligible for conventional DBS therapy. Patients 1–4 (Table 1) were enrolled in a study using an investigational bidirectional neural stimulator ^45^ (Summit™ RC+S model B35300R, Medtronic, Inc.) to develop and test aDBS therapies targeting motor and gait functions (clinicaltrials.gov ID NCT04675398; Fig. 1a). Patient 5 crossed over from a parallel trial (clinicaltrials.gov ID NCT03582891) using the same device.

**Table 1:** Patient demographics.

| Demographics | Patient 1 | Patient 2 | Patient 3 | Patient 4 | Patient 5 |
| --- | --- | --- | --- | --- | --- |
| Sex/Age | M/ in their 60’s | M/ in their 60’s | M/ in their 60’s | F/ in there 60’s | M/in there 50’s |
| Disease Duration (yrs) | 21 | 12 | 8 | 4 | 18 |
| LEDD<sup>a</sup> (mg) | 2590 | 1514 | 1025 | 618 | 1353 |
| **Clinical Measurements** |  |  |  |  |  |
| MDS-UPDRS III OFF/ON<sup>b</sup> | 37/16 | 42/19 | 31/17 | 21/9 | 39/16 |
| MDS-UPDRS III PIGD subscore OFF/ON<sup>c</sup> | -/- | 10/3 | 3/2 | 2/2 | 1/0 |
| MiniBEST LOW<sup>d</sup>/ON | 13/17 | 17/25 | 22/21 | 19/21 | -/- |

a. Levodopa equivalent daily dose
b. Rigidity and postural stability scores excluded due to remote video examination.
c. Total of UPDRS III items: 3.9-3.11 and 3.13.
d. LOW = low medication state defined as the time period between the end of their current medication cycle to the next. Achieved by delaying next dose during testing.

Patients underwent DBS surgery with implantation of a quadripolar depth electrode (Medtronic™ model 3387) targeting the internal segment of the globus pallidus (GPi) and a quadripolar subdural electrocorticography (ECoG) paddle (Medtronic™ model 0913025) placed over the premotor (PM) and primary motor (M1) cortices (Patients 1–4), or over the sensorimotor cortices (M1 and S1) in Patient 5. Electrodes were implanted bilaterally in Patients 1–3 and 5, and unilaterally in Patient 4 due to predominantly unilateral tremor and leg dragging (Fig. 1b). Each Summit RC+S device was connected to a DBS electrode and a subdural cortical paddle from the same brain hemisphere. LFPs and gait kinematic data were recorded during overground walking at self-selected speeds for at least 250 steps, both before stimulation was activated (26–43 days postoperatively), after DBS parameter optimization according to standard clinical care (140–520 days postoperatively), and during aDBS parameter optimization (4-10 sessions). LFPs were recorded from pallidal contacts adjacent to the stimulating electrode and from cortical contacts (PM and M1 or M1 and S1) using adjacent electrode pairs.

Using patient-specific neural data, we developed an aDBS therapy that modulated stimulation amplitude between 0.5x and 1.0x the clinically optimized setting during contralateral leg swing. Control signals were derived from single-frequency band power at one recording site per hemisphere, identified through a data-driven search. Candidate neural features were evaluated based on their ability to distinguish contralateral leg swing from other gait phases. Optimal neural features were embedded into the Summit RC+S device for each brain hemisphere and adaptive parameters manually refined to maximize detection accuracy.

For three patients (Patients 2, 3, and 4), a double-blind crossover experiment was conducted, randomizing three stimulation conditions: cDBS; aDBS ramping from 0.5x to 1.0x their clinically optimized amplitude (ramp-up aDBS, RU-aDBS); and aDBS ramping from 1.0x to 0.5x the clinically optimized amplitude during contralateral leg swing (ramp-down aDBS, RD-aDBS). Each condition was applied continuously for 8–10 days, during which patients filled out a daily motor diary where they rated non-gait related PD symptoms and documented their number of falls and number of freezing episodes (Fig. 1c). Additionally, patients wore a tri-axial accelerometer device (Rover, Sensoplex Inc., Redwood City, CA) to monitor at-home gait metrics.

### Neural biomarkers of leg swing during overground walking

We investigated neural dynamics during natural overground gait by examining LFPs recorded from the pallidum and PM, M1, and S1. Individual gait cycles were extracted offline, and fast Fourier transform spectral power was calculated in the following canonical frequency bands: theta (4–7 Hz), alpha (7–12 Hz), low beta (13–20 Hz), high beta (20–30 Hz), and low gamma (30–50 Hz) bands to assess gait phase dependent modulation. Significant modulation was observed for both ipsilateral and contralateral swing phases in the pallidum and cortical areas, with individual differences in the frequency bands achieving significance (Fig. 2a and Extended Data Fig. 1). Across patients, M1 exhibited the highest proportion of hemispheres with at least one discriminative canonical frequency band (8/9), followed by the pallidum (5/9), PM (3/9), and S1 (2/9) (Extended Data Fig. 1). In three patients, pallidal recordings did not result in any frequency bands capable of distinguishing swing phases.

**Figure 2.**
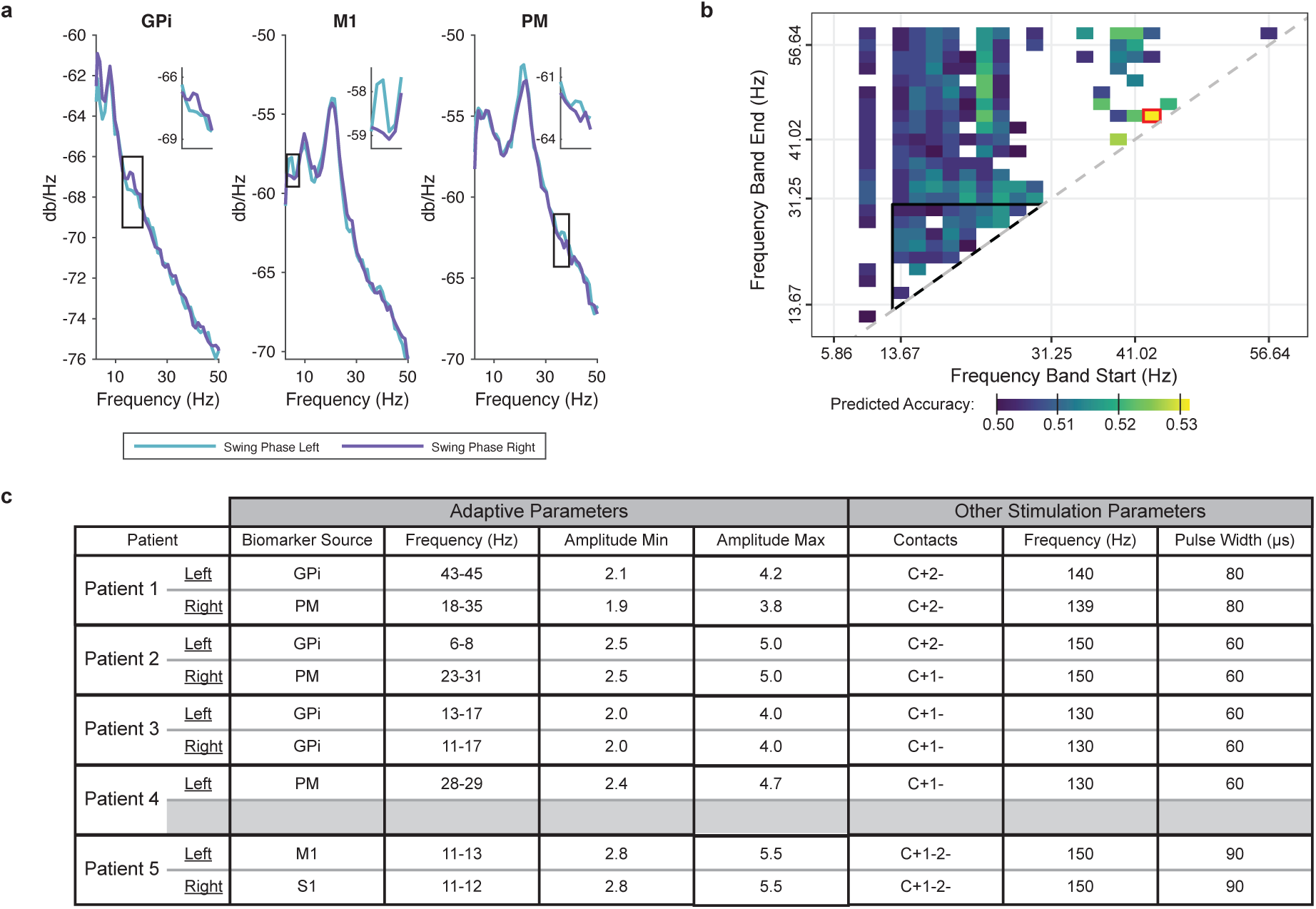
Identification of gait-phase–specific neural biomarkers using cortical and subcortical recordings. **a,** Example power spectral density plots of average values during left and right swing phase in the GPi – left hemisphere, M1 – right hemisphere, and PM – right hemisphere areas. Detectable power differences are observed indicating each area’s potential to find a candidate gait-phase biomarker. Not all patients exhibited detectable power differences in all recording locations. Individual variations between canonical frequency bands are provided in Extended Data Fig. 1. **b,** Example brute-force search results from Patient 1’s left hemisphere for contralateral swing-phase-specific frequency bands using Summit RC+S power calculations. Bands achieving at least 50% accuracy are colored; bands below 50% accuracy are omitted. Many bands within the beta range (outlined by a black triangle) were identified, although the optimal band (42–44 Hz) was outside canonical beta and is highlighted in red. **c,** Table of patient-specific biomarkers to detect contralateral leg swing phase.

Given the observed inter- and intra-patient variability, we next implemented a data-driven approach to identify patient-specific biomarkers of contralateral swing phase that has higher discriminatory power. This approach involved a grid search across all recording locations, evaluating all possible frequency band combinations between 2.5 and 50 Hz, and quantifying contralateral swing-phase decoding accuracy (Fig. 2b). Compared to canonical bands, these individualized frequency bands revealed sub-frequency ranges that better distinguished contralateral swing phase and, in most cases, achieved higher decoding accuracy (Fig. 2b, red square). The origin of patient-specific biomarkers differed between patients and even between hemispheres of the same patient. Optimal gait-phase biomarkers were identified from the pallidum in four hemispheres, the PM in three hemispheres, and M1 and S1 with one hemisphere each (Fig. 2c).

### Accuracy and stability of contralateral leg swing biomarkers in real-time adaptive DBS control

Patient-specific contralateral swing phase biomarkers were embedded in each patient’s neural stimulator using the on-device linear discriminant analysis (LDA) classifier, after which adaptive parameters were optimized. Two parameters were critical: ramping rate and amplifier blanking. The ramping rate, which determines the speed of stimulation amplitude changes after a state transition, was titrated for each patient from 1.9 mA/400 ms upward until either side effects emerged or biomarker accuracy declined due to rapid amplitude changes. Amplifier blanking, which is the brief deactivation of the on-board recording mechanism^45^, was set to 1 ms which was sufficient to minimize ramping-related artifacts.

In real-time testing, the stimulators accurately detected sub-second gait phase transitions: during contralateral leg swing, the binary classifier switched from state 0 to state 1, returning to state 0 during other gait phases (Fig. 3a, Supplementary Information Video 1). The stimulator is able to rapidly ramp up and down stimulation amplitude from 0.5x to 1.0x of the patient’s clinically optimized amplitude during contralateral leg swing. Detection errors occasionally occurred, resulting in shortened or prolonged periods in state 1. Across patients, stimulation amplitude was significantly increased during contralateral leg swing relative to chance, defined as the proportion of the gait cycle spent in swing phase, with an average increase of 18.6% (Fig. 3b).

**Figure 3.**
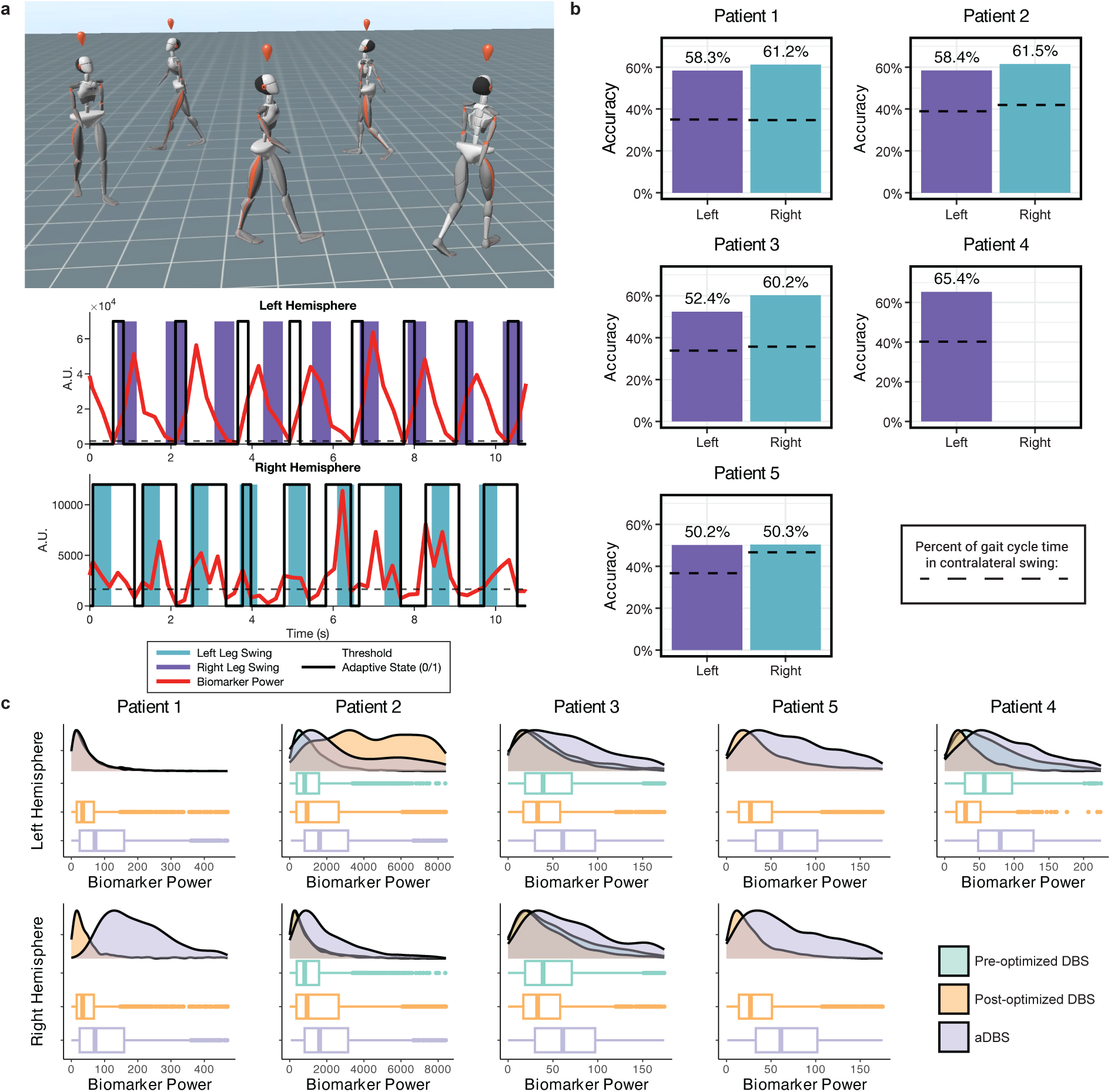
Real-time performance and stability of rapid gait-phase–adaptive DBS. **a,** Example of aDBS implementation during overground walking at a self-selected speed. Spectral power of the selected biomarker (red trace), determined through a data-driven approach, increased above the threshold (dashed black line) during contralateral leg swing (purple, left; teal, right) and decreased below the threshold during other gait phases. When spectral power exceeded the threshold, the DBS system switched to “state 1,” increasing stimulation amplitude (solid black line); when power fell below the threshold, the system returned to “state 0,” decreasing stimulation amplitude. **b,** Accuracy of optimized biomarkers across patients exceeded chance level, with an average improvement of 18.6%. Accuracy was defined using Equation (2) as the ratio of the Summit RC+S state duration in true positive (in contralateral swing phase) plus the duration in true negatives (not in contralateral swing phase) to the total recording duration. Chance level (dashed black line) was determined by the mean proportion of time spent in contralateral leg swing. **c,** Patient-specific aDBS biomarkers tracked across three time points (pre-optimized DBS, post-optimized DBS, and aDBS testing) demonstrated 71.9–98.4% overlap in 12 of 19 comparisons, indicating negligible to moderate differences. Five comparisons showed large differences, each confined to a single hemisphere. Power differences were assessed using Cohen’s D effect size; group comparisons were evaluated by Kruskal–Wallis test followed by Dunn’s post hoc test with Holm–Bonferroni correction.

Biomarker stability was assessed by comparing the spectral power of each patient’s gait-phase specific frequency band across clinical DBS optimization and aDBS testing phases and evaluating the differences in power value distribution. Significant differences in aDBS biomarker power were observed between stimulation conditions in all patients except for the left hemisphere of Patient 1 (Fig. 3c). However, effect size analysis revealed mostly negligible to moderate differences: among 19 comparisons, two were negligible, five were small, seven were moderate, and five were large^46^. Negligible to small differences (Cohen’s D = 0.0398–0.428) corresponded to an 83.4–98.4% overlap in power values^47^. Moderate differences (Cohen’s D = 0.591–0.724) corresponded to a 71.9–76.8% overlap. Large effects were observed in four patients but were limited to one hemisphere each. Overall, patient biomarker spectral power varied modestly across time points, with a high degree of overlap between distributions. Gait parameter changes: in-clinic aDBS vs cDBS

The acute effects of aDBS on gait were assessed against each patient’s clinically optimized cDBS setting. Patients arrived on cDBS and were then switched to individualized aDBS parameters, in which stimulation amplitude ramped from 0.5x to 1.0x the clinically optimized value during swing phase. After 12–15 minutes on aDBS, patients completed 250 overground steps across two recordings.

At the group level, aDBS improved gait symmetry relative to cDBS. Median step length asymmetry decreased by 3.5% with a 3.1% reduction in variability, while median step time asymmetry changed minimally (+0.29%) but showed a 2.4% reduction in variability (Fig. 4a–b). Analysis of step metrics variability showed bilateral reductions in median step length variability of 39.1% (left) and 31.6% (right), and step time variability reductions of 16.0% (left) and 26.6% (right), although these changes did not reach significance (Fig. 4c–d).

**Figure 4.**
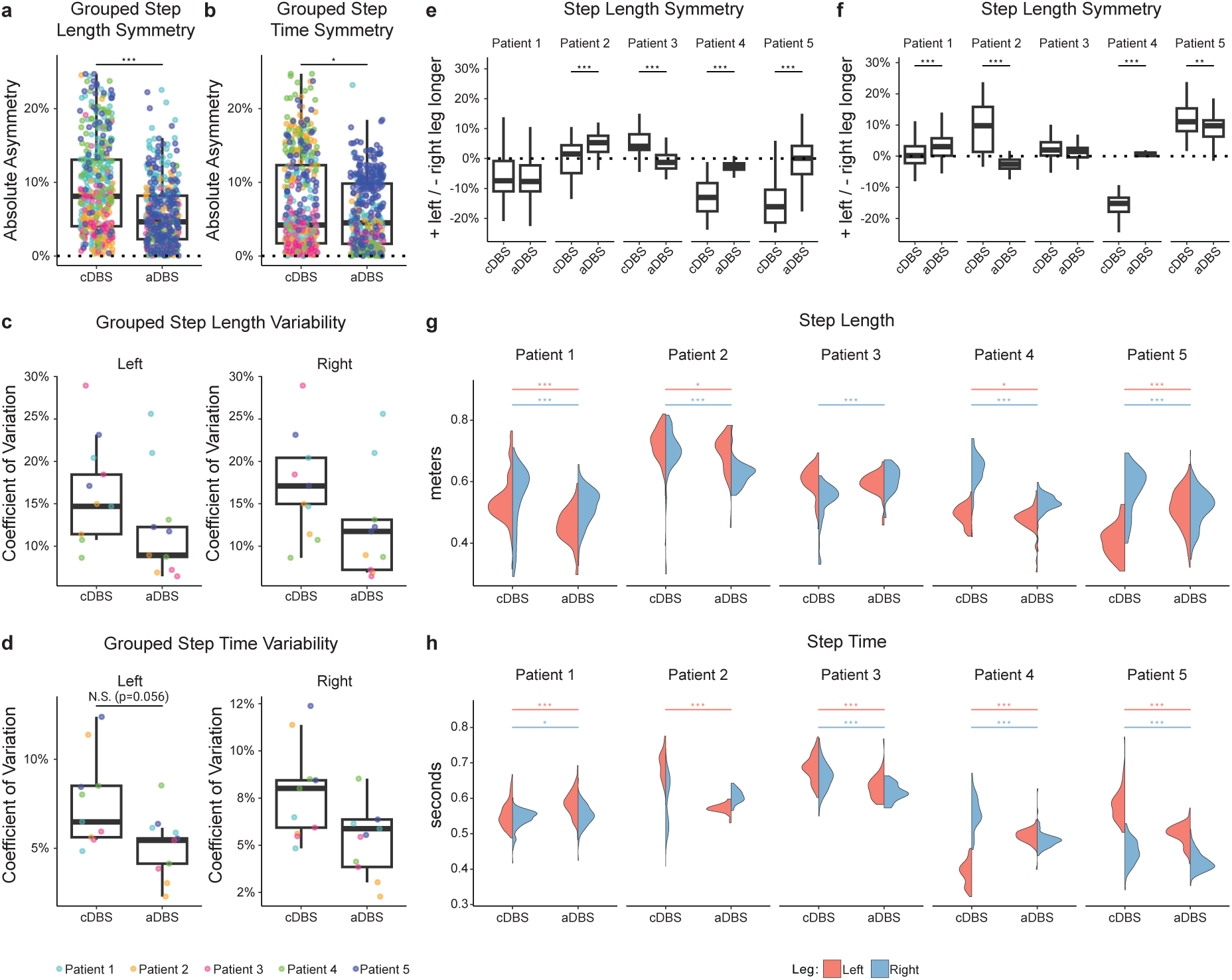
Gait metric changes with adaptive DBS compared to clinically optimized DBS. **a,b,** Group level analysis of absolute asymmetry values, show that aDBS significantly reduced step length (a) and step time (b) asymmetry. Significance levels corresponded to p<0.05 (*), p<0.001(**), and p<0.0001 (***). **c,d,** Step length (c) and step time (d) coefficient of variation (CV) showed consistent reduction under aDBS. Step length CV reduced between 31.6-39.1%, while step time CV reduced between 16.0-26.6%. **e, f,** Step symmetry was calculated as the ratio of the difference between left and right metrics to their sum. Positive values indicate longer or longer-timed left steps; negative values indicate the reverse. Perfect symmetry is 0% (dashed black line). Step length symmetry (e) improved in three patients, whereas Patient 2 exhibited worsened step length symmetry but improved interquartile range (IQR); Patient 1 showed worsened step time symmetry. **g, h,** Violin plots showing changes in step length (g) and step time (h) for the left (red) and right (blue) legs across patients. Step length responses varied among patients, with bilateral decreases (Patients 1, 2, and 4), bilateral increases (Patient 3), and mixed responses (Patient 5). Step time responses were variable and did not consistently mirror step length changes within patients. Changes from clinically optimized DBS to aDBS were assessed using Wilcoxon rank-sum tests. Significance levels are denoted above each plot, with color indicating the leg and asterisks corresponding to p<0.05 (*), p<0.001(**), and p<0.0001 (***).

On an individual level, patient responses varied from one another. Three patients demonstrated significant improvements in step length symmetry, shifting medians toward 0%, indicative of more balanced left–right step lengths and reducing variability by an average of 2.9% (Fig. 4e). Patient 2 exhibited worsened median symmetry (+1% to +5%) but improved variability by 3.9%, while Patient 1 showed no significant change. For step time symmetry, three patients improved significantly, Patient 1 worsened due to prolonged left-leg step time, and Patient 3 showed a small, non-significant improvement (−0.47%, p = 0.132) (Fig. 4f). Step length variability decreased bilaterally in three patients and unilaterally in one, with an average reduction of 60.8% (left) and 78.4% (right), whereas step time variability decreased bilaterally in four patients by 62.9% (left) and 55.6% (right). As with step length variability, Patient 1 exhibited bilateral increases in both step length (+30.6%) and step time variability (+26.6%).

Despite these symmetry and variability improvements, absolute step length and time changes varied between limbs. Significant bilateral step length decreases occurred in three patients (Patients 1, 2, and 4), while two showed unilateral decreases (Patients 3 and 5) (Fig. 4g). Decreases ranged from 1–16%, whereas Patient 3 and Patient 5 exhibited large unilateral increases (13% and 26%, respectively). Step time changes did not consistently mirror step length changes. Patients 2 and 4 displayed mixed step time responses with opposite limb directionality, while Patients 3 and 5 showed step time decreases accompanying step length increases (Fig. 4h).

### Double-blinded cross-over trial shows aDBS decreased falls compared to cDBS

To understand the impact of chronic aDBS stimulation on gait and other PD symptoms, we conducted a double-blind, multi-day evaluation comparing two aDBS strategies —ramp-up (RU-aDBS), previously tested acutely, and a new ramp-down (RD-aDBS) paradigm in which stimulation amplitude decreased from 1.0x to 0.5x during contralateral leg swing—against each patient’s cDBS setting. We hypothesized that ramp up stimulation during contralateral leg swing may suppress excessive pathological oscillations whereas ramp down may permit dynamic, physiological oscillations during this phase of the gait cycle. Three patients (Patients 2, 3, and 4) completed this portion of the study. Patient 1 had undergone an abbreviated test of at-home aDBS due to logistics of living far from our site. Patient 5 had significant disease progression and became wheelchair bound prior to long-term aDBS testing.

Daily motor diaries captured falls, freezing episodes, and self-rated changes in dyskinesia, tremor, and rigidity relative to baseline (“Better,” “Same,” “Worse”), with falls and freezing recorded in categorical bins (0, 1, 2–4, or 5+ events). At the group level, RU-aDBS significantly increased the odds of reporting fewer falls compared to cDBS (odds ratio [OR] = 4.35), while RD-aDBS had no significant effect (Fig. 5a). No group-level changes were detected for freezing episodes. Odds of improvement in rigidity were low under both strategies (RU-aDBS: OR = 0.114; RD-aDBS: OR = 0.038), and no significant effects emerged for dyskinesia or tremor (Fig. 5b).

**Figure 5.**
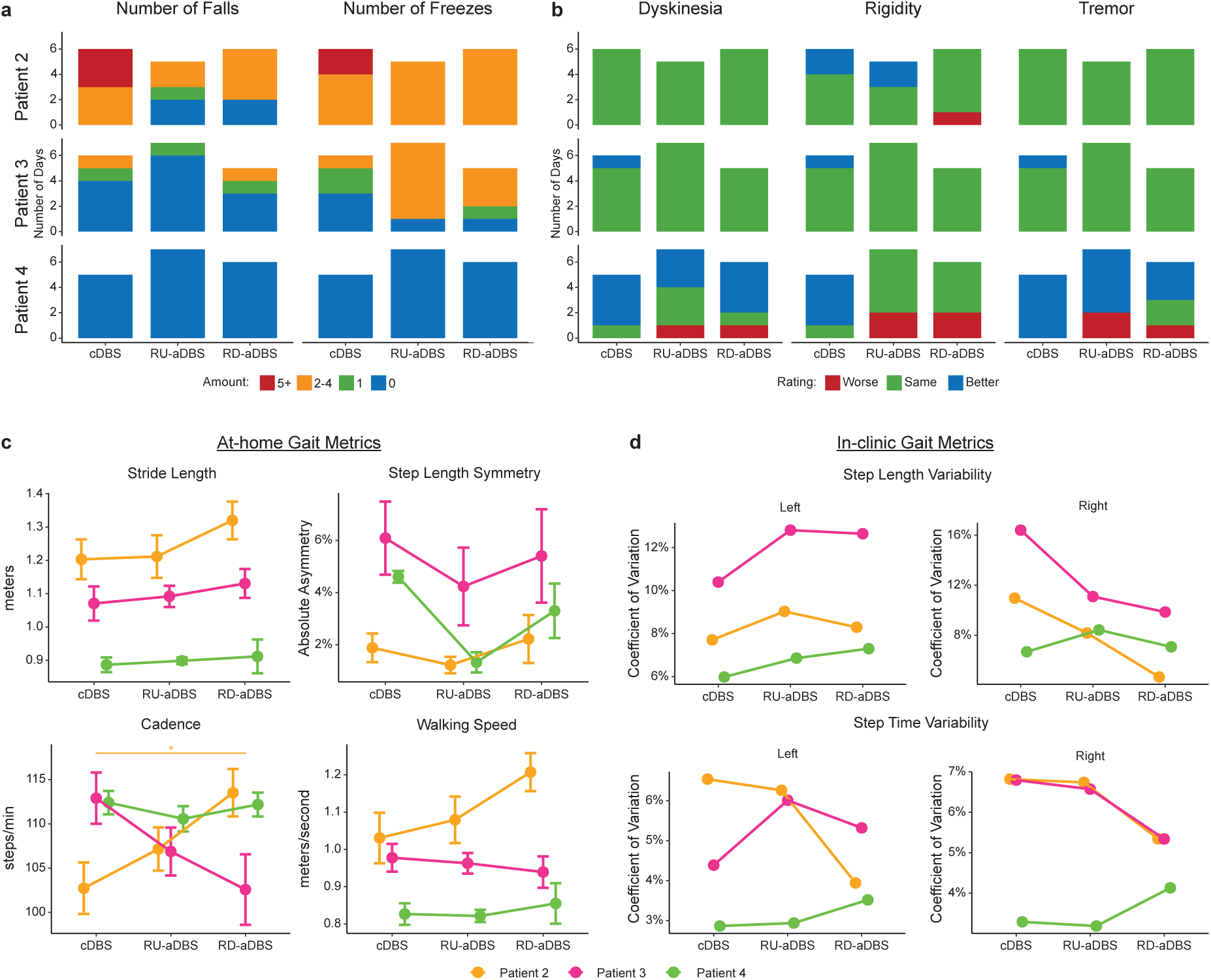
Long term at-home gait metric changes in a randomized double-blinded comparison of cDBS compared to two aDBS settings. Stimulation settings are consistent across all plots: cDBS = clinically optimized DBS; RU-aDBS = aDBS Ramp-Up; RD-aDBS = aDBS Ramp-Down. “Ramp-Up” refers to stimulation amplitude increasing from 0.5x to 1.0x, while “ramp-down” refers to decreasing from 1.0x to 0.5, during during contralateral leg swing. **a,** Stacked bar graphs showing self-reported instances of falls and freezing episodes under each stimulation condition. Bars indicate the number of days they had 0 (blue), 1 (green), 2-4 (orange), or 5+ (red) falls or freezing episodes. RU-aDBS reduced falls for Patients 2 and 3, though it had little effect on freezing for Patient 2 and increased the number of freezing episodes in Patient 3. RD-aDBS had a similar effect, reducing falls while minimally affecting freezing episodes. Patient 4 did not report any falls or freezing under any condition. Group analysis using a cumulative link model indicated that RU-significantly increased the odds of reducing falls (OR = 4.35; *p = 0.04*) **b,** Stacked bar graphs illustrating subjective ratings of dyskinesia, rigidity, and tremor. Bars show the number of days each symptom was rated as “Better” (blue), “Same” (green), or “Worse” (red) compared to their baseline under continuous clinically optimized DBS prior to entering Phase 4. aDBS had little subjective impact for Patients 2 and 3, with most days rated “Same” and only one instance of worsening rigidity (Patient 2). Patient 4 had a mixed response, including several “Worse” and ≥2 “Better” days. A cumulative link model revealed that rigidity had a significantly higher likelihood of being rated “Worse” under both RU (OR = 0.114; *p = 0.017*) and RD (OR = 0.038; *p = 0.0025*) aDBS. **c,** At-home gait metrics collected from a wearable tri-axial ankle accelerometer (Rover) show both RD-aDBS and RU-aDBS improved stride length and step symmetry relative to cDBS. Stride length gains were greater with RD-aDBS (4.7%) than RU-aDBS (0.9%), whereas step asymmetry reductions were larger with RU-aDBS (2.37%) than RD-aDBS (0.36%). Cadence and walking speed changes were minimal (0.51–1.41 steps/min; 0.01–0.04 m/s), although Patient 2 showed a significant (*p = 0.045*) increase in cadence compared with cDBS (+10.79 steps/min). **d,** Both RU-aDBS and RD-aDBS reduced right step length variability (CV, 2–4%) while increasing left step length variability (1.4–1.5%). RD-aDBS decreased step time variability bilaterally, whereas RU-aDBS produced opposing leg-specific effects.

Individual-level responses revealed differences in patients. Patient 2 reported fewer falls under both aDBS strategies, with the largest reduction during RU-aDBS, and a decrease in freezing episodes, though these remained frequent (2–4 per day). Patient 3, who rarely fell under cDBS, experienced further reductions in fall frequency during RU-aDBS relative to RD-aDBS, but freezing episodes increased under both strategies, most prominently with RU-aDBS. Patient 4 reported no falls or freezing episodes in any condition. Symptom ratings mirrored this variability: Patients 2 and 3 reported stable daily scores across dyskinesia, tremor, and rigidity under all conditions, whereas Patient 4 noted consistent improvement under cDBS but mixed responses to aDBS.

Throughout the blinded aDBS testing period, all patient’s at-home gait metrics were captured and analyzed using customized analysis software from wearable tri-axis accelerometers worn on their ankles (Rover, Sensoplex Inc., Redwood City, CA), which computed validated gait measurements^48^, including patient’s stride length (the distance from traveled from one heel strike to the next), walking speed, cadence, and step symmetry. On a group level, both aDBS conditions improved stride length and step symmetry compared to cDBS. For stride length, a larger effect was seen with RD-aDBS (4.7%) than RU-aDBS (0.9%) (Fig. 5c). For step asymmetry, a RU-aDBS showed a greater reduction (2.37%) than RD-aDBS (0.36%). Cadence and walking speed were minimally affected by aDBS compared to cDBS, 0.51-1.41 steps/min and 0.01-0.04 m/s respectively. However, Patient 2 exhibited a marked and significant cadence increase, averaging +10.79 steps/min compared to their cDBS settings.

On the last day of each stimulation condition, gait metrics were calculated from in-clinic testing data. Both RU-aDBS and RD-aDBS consistently improved gait symmetry, with the largest improvements in step length (2.3–2.9%) and smaller but consistent reductions in step time asymmetry (0.7–0.8%) (Extended Data Fig. 2). Both aDBS strategies also reduced right step length variability (2–4%) while increasing left step length variability (1.4–1.5%) (Fig. 5d). Step time variability changes were smaller and more variable, with RD-aDBS inducing bilateral reductions in step time variability, whereas RU-aDBS elicited opposing effects between legs.). Step length and time values showed patient-specific patterns, including bilateral and unilateral changes, opposing leg effects, and difference between step length and step time effects (Extended Data Fig. 2)

## Discussion

Improving advanced gait disturbances with traditional DBS has remained a major clinical challenge. Here, we developed and implemented a novel gait-phase synchronized aDBS therapy that rapidly modulates stimulation amplitude during contralateral leg swing on a sub-second time scale. Personalized neural biomarkers, identified for each hemisphere, served as control signals for aDBS. These individualized biomarkers outperformed canonical frequency bands and demonstrated stability across testing sessions. Acute in-clinic testing revealed that aDBS reduced step length and step time variability and improved gait symmetry. Longer term at-home testing not only confirmed aDBS’s tolerability and maintenance of motor symptom control, but its efficacy for reducing falls and improving step length, cadence, and symmetry compared to continuous DBS.

In this study, we introduce a gait-phase-synchronized aDBS paradigm that achieves sub-second, movement-locked modulation of stimulation amplitude, driven by individualized neural biomarkers derived from cortical–pallidal circuits. This represents a significant technical advance over prior aDBS approaches, which have largely operated on slower time scales (seconds to minutes)^40,49^ and targeted canonical beta activity^41–43,50–52^ without consideration of the movement phase. Given the highly dynamic nature of gait, the first feat was to identify reliable neural biomarkers of contralateral leg swing for each brain hemisphere. Instead of focusing on canonical frequency bands such as beta^41–43,50–52^, which is suppressed during voluntary and repetitive movement^53–55^ and therefore may lack specificity for gait, we adopted a data-driven approach to select patient-specific biomarkers. Interestingly, most personalized biomarkers fell within or spanned canonical frequency ranges but showed superior performance for detecting leg swing compared to traditional bands (Fig. 2c).

The location of the best neural biomarker varied greatly between patients and also between the two brain hemispheres of patients, highlighting the need for multi-focal network sensing that is necessary to implement our aDBS program. By embedding patient-specific spectral features into a chronically implanted bidirectional neurostimulator, we achieved a second unique technical challenging-accurate, real-time detection of contralateral leg swing and rapid ramping of stimulation amplitude within 100–500 ms, precisely timed to the kinematic demands of each step. Previous aDBS implementations typically operated over seconds, with slower ramping to avoid stimulation-induced feedback loops^56^. Despite these challenges, our biomarkers remained accurate and stable, demonstrating the feasibility of rapid aDBS without feedback instability. Our findings demonstrate that this level of control is both feasible and well tolerated in freely moving individuals with PD. To our knowledge, this represents the first implementation of fast aDBS synchronized to a movement event.

Beyond the engineering achievement, our results provide novel insights into the circuit dynamics underlying gait control. We recorded robust, gait-phase-locked modulations across a broad range of frequencies—from theta and alpha to beta and low gamma—in both the pallidal and motor cortical areas. These observations extend previous STN- and cortex-based studies by implicating the pallidum as an active participant in phase-specific gait control, in synchrony with cortical rhythms. The heterogeneity in optimal biomarkers across patients suggests that gait control relies on individualized patterns of cortico-basal ganglia coordination, shaped by disease progression, medication state, and perhaps individual differences in gait strategies.

From a systems neuroscience perspective, the convergence of low-frequency rhythms linked to bilateral coordination and higher-frequency activity associated with motor execution supports a multi-band, distributed control model of locomotion. Recent evidence increasingly supports the view that gait is represented within distributed cortico-basal ganglia circuits. Recordings from the subthalamic nucleus (STN) and cortex in individuals with PD have demonstrated gait-phase–locked modulations in LFPs, particularly in low-frequency bands including theta, alpha, and beta^31,32,35,57,58^. These studies suggest that basal ganglia and cortical networks are dynamically engaged throughout the gait cycle, with distinct spectral modulations occurring during specific gait phases. Our findings align with and extend these observations. We recorded robust gait-phase-locked modulations in both the GPi and motor cortex during natural overground walking, with modulations spanning a broad frequency range from theta through low gamma (30–50 Hz). While beta-band activity has historically been emphasized due to its association with bradykinesia and rigidity, we observed that multiple frequency bands— including theta, alpha, beta, and low gamma—were modulated during different phases of the gait cycle. In particular, lower-frequency rhythms such as theta and alpha, which have been associated with phasic muscle activation during locomotion^59,60^, may play a critical role in coordinating bilateral limb movements and maintaining rhythmic gait patterns. Given the need for precisely timed alternation of limb movements and balance adjustments during gait, lower-frequency synchronization across cortico-basal ganglia circuits may contribute to the integration of bilateral motor outputs, while higher-frequency modulations may reflect more localized aspects of motor execution. Personalized biomarkers derived from these frequency ranges were highly effective in detecting contralateral leg swing and driving gait-phase-synchronized aDBS, further supporting their functional relevance. Future studies incorporating bilateral cortical and subcortical recordings will be important to delineate how different frequency bands cooperate across brain regions to support coordinated walking, and how disruptions in these multi-band rhythms contribute to gait impairments in PD.

Clinically, gait-phase-synchronized aDBS produced improvements in step length and step time variability and bilateral symmetry—metrics that are closely tied to instability and fall risks^61–63^ and rarely improved with cDBS^64,65^. The mechanistic implication is that continuous stimulation, while beneficial for certain cardinal symptoms, may interfere with the physiological timing signals needed for dynamic balance and limb coordination. The “information lesion” hypothesis posits that continuous DBS alleviates symptoms by disrupting pathological network communication^66–68^. Suppression of beta activity following DBS supports this concept^69–77^. In the context of gait, this could mean suppression of critical sensory-motor cues that are phase-locked to the walking cycle. Our approach preserves these windows for physiological processing by delivering stimulation only when pathological activity is most likely to impair performance— during specific phases of the gait cycle—while minimizing interference at other times. This temporally selective strategy likely explains why aDBS improved dynamic gait control more than cDBS.

The timing of stimulation may be key to modulating network dynamics. Coordinated reset stimulation, which is designed to desynchronize pathological synchrony, requires precise temporal delivery and has been associated with motor improvements^78–82^. Although high entropy (i.e., desynchronized activity) in basal ganglia activity may worsen parkinsonism^83–85^, excessively low entropy is associated with hyperkinetic disorders^86^, suggesting the existence of an optimal entropy range. We hypothesize that gait-phase-synchronized aDBS introduces semi-random stimulation variability, modulating network entropy to a “sweet spot” that supports effective gait control without destabilizing other motor functions.

A further advance in this work was the integration of long-term, at-home gait monitoring to assess real-world mobility. These data revealed benefits that were not consistently detectable during brief in-clinic testing, highlighting the considerable day-to-day variability in gait performance. Home-based assessment provides a more ecologically valid measure of function and captures the cumulative impact of therapy on fall risk and mobility. Our finding that aDBS may reduce falls in real-world conditions strengthens its translational relevance and points to the value of wearable sensor technology in both clinical trials and long-term patient management.

Our study has limitations. The small sample size reflects the invasive nature of device implantation and the stringent inclusion criteria for gait disturbances. Additionally, while our platform allowed detection based on a single frequency band, future implementations incorporating multiple bands may improve swing phase detection accuracy^87^. Finally, not all patients responded equally. Patient 1, who exhibited severe dyskinesia, showed no improvements in gait metrics, possibly due to the chronic instability of stimulation settings and limited acute testing duration. These findings highlight the need for longer adaptation periods in future studies.

## Conclusion

Our results demonstrate that sub-second, gait-phase-synchronized adaptive DBS—driven by individualized cortical–pallidal biomarkers—can restore dynamic gait control in Parkinson’s disease while preserving broader motor function. By targeting stimulation to the precise temporal windows when pathological activity is most disruptive, this approach overcomes key limitations of continuous DBS and reveals mechanistic insights into the circuit-level control of locomotion. The integration of home-based gait monitoring further establishes its real-world relevance. These findings position movement-state–driven aDBS as a viable and potentially transformative therapy for gait disorders, paving the way toward a new class of precision-timed neuromodulation strategies that aim not just to suppress symptoms, but to re-establish the physiological dynamics of human movement.

## Methods

### Clinical trial overview

This study was completed within the *Adaptive Deep Brain Stimulation to Improve Motor and Gait Functions in Parkinson’s Disease* clinical trial (ClinicalTrials.gov; NCT04675398). The primary outcome of this trial is to deliver personalized neurostimulation based on individual physiological biomarkers to enhance locomotor skills in patients with PD. All data presented here are part of the ongoing exploratory clinical trial and do not contribute towards any conclusions regarding the primary outcome of the trial. The neural device used in this study (Medtronic Summit RC+S) was approved by the Food and Drug Administration as an investigation device exemption, which was followed by study protocol approval by the University of California, San Francisco Institutional Review Board (20-32847). Five patients gave their informed consent to participate in this trial following multiple conversations with study investigators in which the details of study enrollment, including risks related to the study device, were explained to them.

### Patients

We recruited four patients (3 males, 1 female; between 60-70 years old) with Parkinson’s disease who were undergoing deep brain stimulation between June 2021 to October 2022, and one patient (male; in their 50’s) joined our study while they were enrolled in another clinical trial (NCT03582891). Patients exhibited gait impairments, had at least a 30% improvement in MDS-UPDRS III score on antiparkinsonian medication, and had a Montreal Cognitive Assessment score ≥ 21 (Table 1). All patients were recruited from outpatient Neurology or Neurological Surgery clinics at the University of California, San Francisco.

### Neural implants and lead reconstruction

The investigational neural device used in this study (Medtronic Summit RC+S B35300R) is a rechargeable neural interface, capable of simultaneously delivering stimulation to the DBS target and recording neural local field potentials (LFP). Up to four bipolar electrode pairs can be recorded from. Additionally, it is capable of a fully embedded adaptive algorithm.

Four patients (1, 2, 3, and 5) underwent bilateral implantation and one patient (4) underwent left hemisphere unilateral implantation of a cylindrical quadripolar DBS lead into the GPi (Medtronic model 3387) and a quadripolar cortical paddle over the cortex (Medtronic model 0913025). The cortical paddles were positioned such that the electrodes spanned over the M1 and PM cortices in Patients 1-4, while Patient 5 was positioned over M1 and S1. A DBS and cortical paddle lead from the same hemisphere was connected to each Summit RC+S neural stimulator.

Precise electrode localization was performed using established image analysis pipelines for depth and cortical electrodes^29^. Briefly, post-implantation high resolution CT images were coregistered to preoperative T1-weighted 3T MRI using a rigid, linear affine transformation. Coregistration accuracy was verified by visual inspection and, when necessary, refined using an additional brain shift correction routine to align subcortical anatomy. Then, electrode artifacts were identified on the CT images and fit to know electrode geometry. Cortical electrodes were additionally projected to the MRI-rendered pial surface. Lastly, electrode locations were normalized into Montreal Neurological Institute space^88^ and visualized either on the FreeSurfer average cortical surface^89^ or a Parkinson’s disease specific standard subcortical atlas^90^.

### Experimental paradigm

Patients 1-4 were brought into the lab twice prior to adaptive DBS testing. The first research visit occurred between 26-43 days post-implantation prior to turning on DBS, and the second visit occurred once a UCSF neurologist determined their clinically optimized DBS parameters (140-520 days). Patient 5, who joined our study after being clinically optimized, was brought into the lab once prior to adaptive DBS testing. Each visit the patients were instructed to walk overground at their self-selected speed for 250 steps. This was repeated in the ON-medication state, and in a LOW-medication state. A LOW-medication state was defined as 15 minutes prior to their next medication dose. Data from these visits were used to generate patient specific candidate biomarkers to detect the contralateral leg swing.

The biomarkers served as the control signal for our embedded adaptive DBS algorithm, which aimed to change the stimulation amplitude from their clinically optimized stimulation amplitude during contralateral leg swing to half the value during all other gait phases (Fig. 1a). The stimulation frequency, pulse width, and electrode contacts did not change from their clinically optimized values to minimize side effect potential. Each biomarker was evaluated for accuracy by turning on adaptive DBS for 15-20 minutes, having them perform the walking task above, and comparing the output of the DBS system with the timings of the contralateral leg swing. Biomarkers with the highest accuracies were tested again to optimize parameters such as the threshold and how fast the stimulation amplitude would increase or decrease. More details on how these biomarkers were determined, evaluated, and optimized can be found in section *“Adaptive Deep Brain Stimulation”*.

Optimized adaptive DBS settings for Patients 2, 3, and 4 were tested in a double-blind experiment by randomizing three conditions: clinically optimized DBS settings (cDBS); adaptive DBS ramping from 0.5x to 1.0x the clinically optimized amplitude value during contralateral leg swing (RU-aDBS); and adaptive DBS ramping from 1.0x to 0.5x the clinically optimized amplitude value during contralateral leg swing (RD-aDBS). Each condition was tested throughout the patient’s entire waking period and was maintained for 8–10 days. During testing, patients wore ankle accelerometer devices (Rover, Sensoplex Inc, Redwood City, CA), each containing a sensor comprised of a triaxial gyroscope, accelerometer, and magnetometer which recorded kinematic data locally. Patient’s also completed post-test motor diaries in the evening.

### Data acquisition

Gait kinematics were collected from the patients using two wireless systems: the Delsys Trigno/Centro (Delsys Inc. Natick, MA) and Xsens MVN Analyze (Movella, Enschede, The Netherlands). The Delsys system included eight IMU+EMG sensors placed bilaterally on top of their neural implant, hands, tibialis anterior, and soleus; two goniometers (SG110/A) placed next to the lateral malleolus; and eight force sensitive resistors (FSR; DC:F01) placed bilaterally on the toe, first metatarsal, fifth metatarsal, and heel^31^. The Xsens system comprised of 15 IMU sensors and place in accordance to Xsens instructions for wireless motion tracking.

Neural LFPs were recorded from one subcortical and two cortical bipolar electrode pairs. Globus pallidus potentials were recorded from contacts adjacent to the stimulating cathode, allowing for common mode rejection of the stimulation artifact when stimulation was being delivered. Cortical recordings were recorded from adjacent pairs with the anterior pair overlaying the PM cortex. All LFP data was recorded at 500 Hz and passed through a pre-amplifier high-pass filter of 0.85 Hz. The data was then passed through a low-pass filter of 100 Hz for subcortical signals and 450 Hz for cortical signals, amplified, and passed through another low pass filter of 100 Hz for subcortical signals and 1700 Hz for cortical signals. Triaxial accelerometry data was also collected from the Summit RC+S system and was sampled at 64 Hz. All data from the Summit RC+S system was extract and processed using the OpenMind consortium open-source code (https://github.com/openmind-consortium/Analysis-rcs-data)^49,91^.

### Data analysis

Neural LFP data collected prior to adaptive DBS was inspected for stimulation, electrocardiogram (ECG), and nonphysiologic polyphasic artifacts. Stimulation artifacts were filtered using a 4^th^ order Butterworth filter at the patient’s clinically optimized stimulation frequency and at half the value to remove harmonics. ECG artifacts were removed using a template subtraction method by providing a manually selected ECG artifact^92^. In rare cases, a nonphysiologic polyphasic artifact would be present and were removed using the same template subtraction method to remove ECG artifacts.

Gait events were extracted from the toe and heel FSRs using a custom MATLAB script. Heel-strikes were defined as the time when the heel FSR reading was above 5% in the positive direction. Toe-offs were defined as the time when the toe FSR reading was below 5% in the negative direction. In some instances, gait events were estimated using signals from the other intact sensors when toe or heel FSRs were damaged during an overground walking bout.

Filtered neural LFP, gait events, and gait kinematic data were synchronized by aligning the peaks in acceleration captured by the Summit RC+S device(s), Delsys sensors over the Summit RC+S, and Xsens acceleration around the shoulders^31^. Wavelet spectral analysis (MATLAB *cwt* function) was applied to the LFP signals, and the power spectra for each gait cycle was extracted and normalized to the average power during movement. Separately, the output of the Summit RC+S fast Fourier transform (FFT) was simulated using the raw LFP signals using the Python *rcssim* package^93^ (https://github.com/Weill-Neurohub-OPTiMaL/rcs-simulation). Spectral power during the left and right leg swing phase was calculated within the theta (4-7 Hz), alpha (7-12 Hz), beta (13-30 Hz), and low gamma (30-50 Hz), to see if any bands exhibit a power difference.

In-clinic gait metrics were derived from recordings obtained with the Delsys and Xsens systems. Temporal metrics included step time, swing time, and double support time, extracted from gait events detected by FSRs of the Delsys system. Step length was the only spatial metric and was calculated from foot position data recorded by an Xsens sensor placed on top of each patient’s shoe. Step time was defined as the interval from heel strike of one foot to heel strike of the contralateral foot. Step length was defined as the distance from the heel of one foot to the heel of the contralateral foot at heel strike.

Symmetry was calculated for step length and time as the difference between the left and right leg divided by the sum (eq. 1). From this equation, a positive value implies the left leg has a higher value, a negative value implies the right leg has a higher value, and a value of 0 would implies perfect symmetry where the left and right leg values are equal.

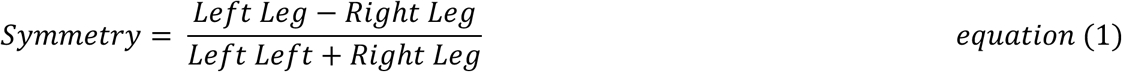

At-home gait metrics were obtained using the Rover wearable device (WD) consisting of a 38.1 mm x 51.2 mm x 13.7 mm, lightweight rechargeable inertial sensor module paired with a fabric strap allowing the device to be worn around the ankle. The 9-axis motion sensor module contains a triaxial gyroscope, triaxial accelerometer, and triaxial magnetometer which sample at 100 Hz and record data locally on a secure digital card. WD data were uploaded by patients to the secure Rover cloud. Once a WD recording was uploaded to the cloud, the Rover Gait Analysis System (GAS) was used to generate a multiple-page gait analysis (MPGA) spreadsheet. Each MPGA summarized activity details (e.g., recording duration, total walking time, total distance walked, etc.) for each recording. In addition, it provided a list of time-stamped strides for both legs with gait cycle time (s), length (cm), swing period (s), and heading (°) recorded. MPGA reports were used to calculate average gait metrics for all patients during the long-term aDBS phase.

### Statistical analysis

Differences in wavelet and simulated Summit RC+S spectral power during the left and right swing phases were tested for each patient, frequency band, hemisphere, and recording location separately. Normality was first tested for each group using the Shapiro-Wilk test, with all tests achieving a p-value < 0.05, indicating that all extracted spectral power data were not normally distributed. Significance testing for each group was performed using the two-sided Wilcoxon rank-sum test.

Biomarker spectral power, computed from the Summit RC+S, for each patient was compared at three different time points, before and after optimization of clinical DBS parameters and during aDBS testing. A Shapiro-Wilk test determined that the spectral power was not normally distributed. Thus, a Kruskal-Wallis test was performed across the three time points. To determine which time points are significantly different from each other, a post-hoc analysis was performed using two-sided Dunn’s test with a Holm-Bonferroni p-value adjustment for multiple comparisons.

Effect size analysis was also performed across the three different time points using Cohen’s D statistical test for each pair of time points. Differences of power between pairs of time points were classified as negligible, small, moderate, and large^46^.

Temporal and spatial gait metrics and symmetry changes from continuous DBS to adaptive DBS setting were tested for each patient. Values were marked as outliers if they were not contained within the 1.5 times the interquartile range and removed. Statistical testing followed the same scheme, where normality was tested first, indicating that these metrics were not normally distributed, and then a two-sided Wilcoxon rank-sum test was performed to compare the two DBS settings.

Patients completed end-of-day motor diaries during Phase 4 to rate the severity of three cardinal Parkinson’s disease symptoms—dyskinesia, rigidity, and tremor—relative to their baseline experience under continuous clinically optimized DBS and recall the number of falls and freezing episodes they experienced during the day. Ratings for the cardinal symptoms were recorded on a 3-point ordinal scale: “Worse,” “Same,” or “Better.” For the number of falls and number of freezing episodes, patients selected one of four options: 0, 1, 2-4, and 5+. To evaluate whether stimulation setting influenced symptom perception, the number of falls, or number of freezing episodes, a cumulative link model was used to assess the probability of reporting a change from baseline across conditions.

### Adaptive deep brain stimulation

The adaptive algorithm was designed to increase or decrease the stimulation amplitude while keeping the stimulation frequency, pulse width, and stimulating electrode contact the same. The stimulation amplitude ranged from 0.5x to 1.0x the patient’s clinically optimized therapeutic value to ensure a baseline level of stimulation was delivered at all times. Changes in stimulation amplitude were targeted to only occur during contralateral leg swing (Fig. 1a).

Embedding the algorithm into the Summit RC+S system involved setting several values: the biomarker frequency band(s), the rate of change for the stimulation amplitude (both increases and decreases), the onboard FFT parameters, the state parameters, and the threshold for changing states^49,56^. Here, “state” refers to a set of stimulation parameters that the Summit RC+S system will adjust based on whether the STFT power of the biomarker is above or below the threshold. Our algorithm used a single frequency band, which required us to set two states and a single threshold value. After setting these values, the biomarker was evaluated for accuracy, and if it showed the highest accuracy, the threshold and rate of change were further optimized.

#### Gait phase biomarker search

A data-driven approach was used to determine which frequency band to test as the biomarker for the adaptive algorithm. First, the Summit RC+S device’s onboard FFT power over time was simulated using the Python *rcssim* package^93^. This package allowed us to simulate different FFT parameters used by the Summit RC+S system, including the number of LFP data points to use in the calculations (256 or 1024) and how often a new FFT spectral power calculation is performed (205, 256, or 410 ms); these values were chosen to comply with the constant-overlap and add property.

After FFT power simulation, arbitrary-length frequency bands were created between 2.5 and 50 Hz, where the power was summed. Labels were added to the frequency bands, marking contralateral leg swing with a “1” and “0” otherwise. For each frequency band, 15 thresholds were calculated between the 15% quantile and the 85% quantile in increments of 5% of the power observed in that frequency band. Finally, the thresholds were used to calculate the accuracy of the settings. The 20 biomarkers with the highest accuracies were then selected to be tested with the patients.

#### Biomarker evaluation

Candidate biomarkers were embedded into the patients’ left and right Summit RC+S systems and tested one at a time. Patients sat in a chair with armrests for 1 minute after adaptive DBS was activated, during which they informed the research team of any side effects. If a side effect did not subside within the next minute, the biomarker trial was stopped, the patient was returned to their clinically optimized setting, and the next biomarker was tested. Across all patients, three side effects were reported: Patient 1 experienced dyskinesia primarily affecting the right side of their body, Patient 3 experienced visual abnormalities, and Patient 4 experienced muscle pulling around the right jaw. Once the patients were returned to their clinical DBS settings, the side effects ceased.

Biomarker settings that did not cause side effects continued testing with 1 minute of standing arm-swing and overground walking at a self-selected speed for 250 steps. During the overground walking portion, a licensed physical therapist (J.E.B) guarded the patient to prevent any falls. Overground walking data was then processed offline and analyzed for accurate changes in stimulation amplitude during the contralateral swing phase. Accuracy was calculated by summing the total time the patient spent in contralateral leg swing (detected by the Summit RC+S system as a true positive) and the total time spent in other gait phases (when the Summit RC+S system correctly did not detect swing phase as a true negative). This sum was then divided by the total length of the trial (eq. 2).

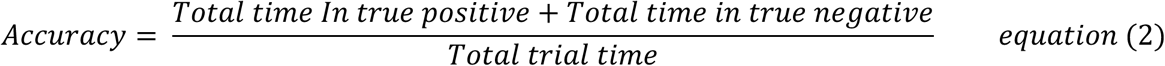

#### Biomarker optimization

The biomarkers that achieved the highest accuracy for the left and right Summit RC+S systems were tested in the next adaptive testing session to optimize the threshold, the rate of change of the stimulation amplitude, and the state parameters. Thresholds were increased or decreased based on visual inspection of the Summit RC+S-reported stimulation amplitude during overground walking. Across all patients, threshold adjustments were within ±10% of the original threshold predicted by the data-driven simulation. The rate of change for the stimulation amplitude was initially set to 1.9–2.7 mA/400 ms, depending on the patient’s clinically optimized value, and was reduced if side effects were observed. Lastly, the two states were switched if the amplitude decreased during the contralateral leg swing.

#### Double-blind Testing

The optimized adaptive setting, referred to as aDBS Ramp-Up, was evaluated in a double-blind study against each patient’s clinically optimized setting and an additional adaptive setting, aDBS Ramp-Down. The Ramp-Down protocol reversed the Ramp-Up stimulation pattern by decreasing amplitude from 1.0x to 0.5x during contralateral leg swing. Each setting was tested over 8–10 full days. At the end of each day, patients completed a motor diary rating dyskinesia, rigidity and tremor relative to their baseline under continuous DBS. They also recorded the number of falls and freezing episodes experienced. On the final day of each setting, patients returned to the clinic for an in-person research assessment.

## Data availability

Data collected in this study is available under restricted access, in compliance with our clinical trial protocol. Upon reasonable request, investigators outside UCSF may receive de-identified data, but it must not be shared with others without prior approval. Data access requests can be made to Dr. Doris Wang. The source data used to generate the figures in this study will be made available with all patients’ data de-identified.

## Code availability

Except for the biomarker identification code, all MATLAB, Python, and R analysis code used to analyze and generate the main findings of this study will be made available at (https://github.com/UCSF-wang-lab/gp-FaDBS) within one month of publication. The biomarker identification code will be made available after subsequent manuscripts that use this code are published. Code written to interface with and process raw data from the Medtronic Summit RC+S is available in the OpenMind public GitHub repository (https://openmind-consortium.github.io).

## Acknowledgements

This study was supported by the Michael J Fox Foundation Grant MNS135499A, the UCSF Burroughs Wellcome Fund Career Award for Medical Scientist, and National Institute of Neurological Disorders and Stroke (NIH/NINDS) 1R01NS130183. This study was also partially supported by UCSF Catalyst Grants. All funding above was obtained by D.D.W. The OpenMind consortium, which provided software to interact with the investigational device and process the raw data, was funded by NINDS U24 NS113637 (to P.A.S). We thank T. Wozny for lead localization (Fig. 1b), R. Gilron, C. Smyth, and M. Shcherbakova for technical contributions, and N. Sirivansanti and Ken Probst for medical art (Fig. 1a). Lastly, we thank the patients and their spouses for their involvement in the study.

## Author information

### Contributions

K.H.L and D.D.W designed the study and analysis pipeline. K.H.L, J.P.B, J.E.B, S.S, H.F.A, and J.H.M collected the data. K.H.L analyzed the data. J.P.B and J.H.M facilitated patient communication and coordination throughout the study. K.H.L and D.D.W drafted the manuscript, and all authors reviewed, commented, and approved the final version.

## Ethics declaration

D.D.W. consults for Medtronic, Boston Scientific, and Iota Bioscience, and research support from Boston Scientific. P.A.S. receives support from Medtronic and Boston Scientific for fellowship education. K.H.L, J.P.B., J.E.B., S.S, J.H.M., H.F.A., and J.T.C declare no competing interests.

## Additional information

### Supplementary information

A supplementary video of Fig. 3A is provided.

## Extended Data

**Extended Data Figure 1.**
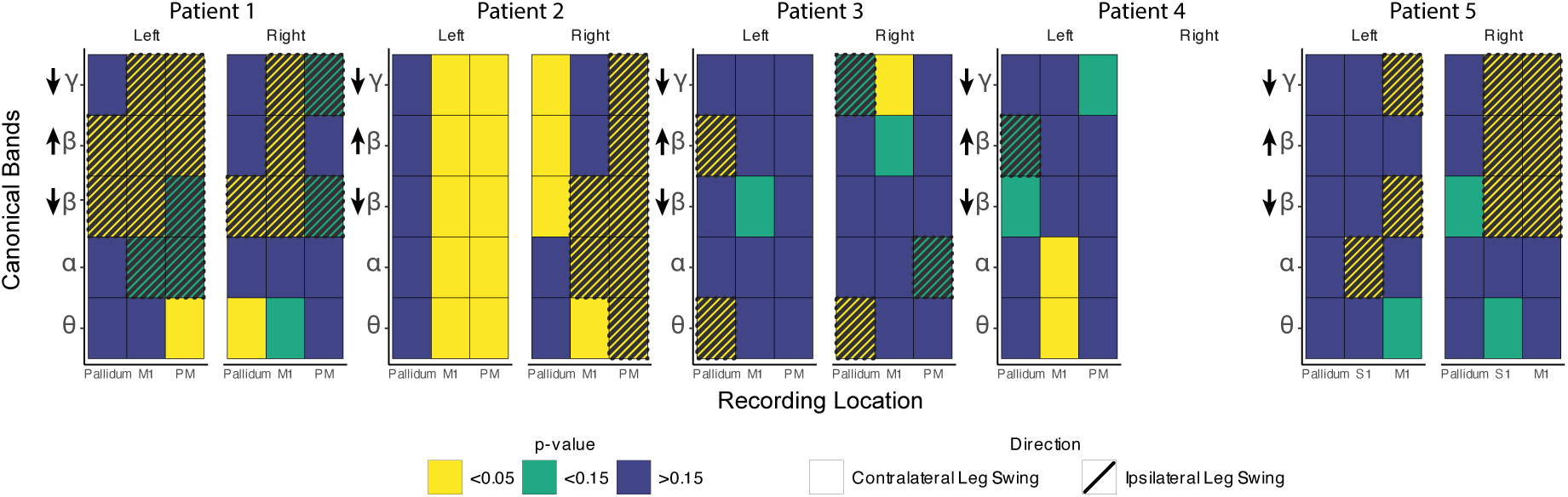
Differences in canonical frequency band powers in distinguishing leg swings in individual patients. Heatmap of canonical band significance test for distinguishing between ipsilateral or contralateral leg swing phases compared to the entire walking period. Color indicates significance level: p<0.05 (yellow), p<0.15 (green), p>0.15(blue). Frequency bands: Θ = 4-7 Hz, α = 7-12 Hz, ↓β = 13-20 Hz, ↑ β = 20-30 Hz, ↓γ = 30-50 Hz. Recording location: Pallidum = GPi, M1 = primary motor cortex, PM = premotor cortex, S1 = primary somatosensory cortex.

**Extended Data Figure 2.**
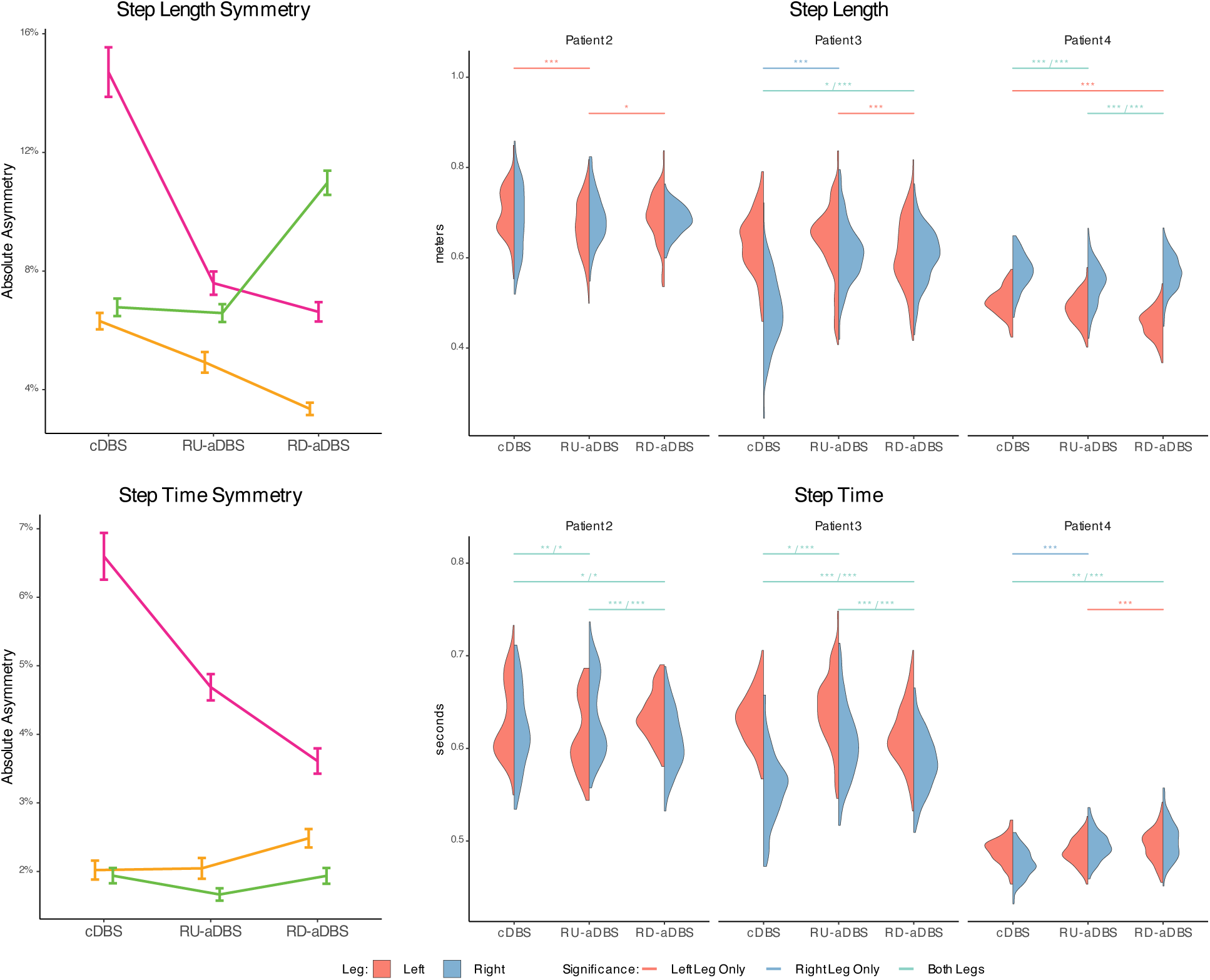
Long term in-clinic gait metrics. Double-blind testing in-clinic gait metrics. Violin plot color indicates left (red) or right (blue) leg. Significance level color indicates whether left (red), right (blue), or both (green) legs were significantly different. Significance levels are p<0.0005 (***), p<0.005 (**), and p<0.05 (*).

